# Individual cardiorespiratory fitness exercise prescription in elderly based on BP neural network

**DOI:** 10.1101/2022.04.06.22273528

**Authors:** Yiran Xiao, Chunyan Xu, Lantian Zhang, Xiaozhen Ding

## Abstract

Cardiorespiratory fitness (CRF) declines as age increases in elderly. An individualized CRF exercise prescription can maintain the CRF level and delay aging process. Traditional exercise prescriptions are general and lack of individualization. In this paper, a new study based on back-propagation (BP) neural network, is investigated to predict the individualized CRF exercise prescriptions for elderly by correlate variables (age, sex, BMI, VO_2max_ initial value, improvement etc.). The raw data are split to two parts, 90% for training the machine and the remaining 10% for testing the performance. Based on a database with 2078 people, the exercise prescription prediction model’s MAE, RMSE and R^2^ are1.5206,1.4383 and 0.9944. 26 female subjects aged 60-79 years are recruited to test the model’s validity. The VO_2max_’s expected improvement was set at 10%. Based on the basic information of these elder women, we get personalized exercise prescription (frequency, intensity, time and volume) of each subject. All of them finished their own exercise intervention. The results show that the post VO_2max_ was significantly different from the pre VO_2max_ and improved by 10.1%, and a total of 20 subjects(74.1%) improved within one standard deviation and 25 subjects(92.6%)improved within 1.96 times standard deviations. Our study shows that a high degree of accuracy in exercise suggestions for elderly was achieved by applying the BP neural network model.

## Introduction

Cardiorespiratory fitness (CRF), also known as Cardiovascular fitness, is a health-related component of physical fitness which refers to the capacity of the circulatory respiratory systems to supply oxygen to muscular systems during physical activity[1]. Longitudinal studies have found that the decline of CRF over time range from 5% to 20% per decade from the age of 30 onwards[2], with prior studies demonstrating a greater rate of decline in older age[3,4]. CRF is associated with cardiovascular disease (CVD) and all-cause mortality in both men and women[5]. In addition, specifically among the elderly, satisfactory CRF is also required for quality of life, preservation of function, and independence[2–6].

In all methods to improve the CRF of elderly, according to given exercise prescription, supervised exercise training is one of the well-established strategies. Exercise prescription is a specific plan of fitness-related activities designed for a specific purpose (such as maintaining CRF), which is usually developed by a fitness or a rehabilitation specialist. It mainly includes the exercise frequency, intensity, type, and time (FITT), also includes volume, and progress (VP). Currently, the WHO recommends that the elderly should do at least 150-300 minutes of moderate-intensity aerobic physical exercise; or at least 75-150 minutes of vigorous aerobic physical exercise; or an equivalent combination of moderate-intensity and vigorous exercise throughout the week, for substantial benefits to CRF[7]. However, there is substantial heterogeneity in CRF response to a certain exercise prescription; some participants got a high improvement in CRF levels, some had no improvement with training[8,9]. In these studies, it may be limited by the high heterogeneity of dose parameters, participant characteristics, or both[10]. The ACSM standpoint to ensure improved CRF in older adults recommends at least 30 minutes of moderate-intensity exercise at least 5 days per week or at least 20 minutes of high-intensity exercise at least 3 days per week. Furthermore, in previous studies have adopted a variety of methods to formulate an individualized CRF exercise prescription for elderly[11], For example, after evaluating the basic physical condition of elderly, the doctor formulates the exercise prescription for elderly people according to their own experience, but this is not individualized exercise prescription, because of the possibility of directly adopting predecessors’ exercise prescription, and often fail to achieve the desired improvement effect[12]. Therefore, factors such as age, sex, and physical fitness need to be considered when formulating personalized exercise prescriptions to improve CRF in the elderly.

Therefore, it is necessary to collect the basic information of subjects in advance to achieve accurate and individualized exercise prescription prediction. At present, there are many methods for prediction, such as traditional prediction method, regression analysis method and gray theory prediction method etc. The disadvantages of these methods in practical applications include: (1) Due to nonlinear prediction model, the parameters are difficult to determine, (2) Forecast factors obtained from relevant analysis are often fuzzy and uncertain, so prediction accuracy is low and it’s hard to get ideal results[13].

In order to solve this problem, we propose a novel model based on BP neural network that has a good ability of deep learning and inferencing. For nonlinear systems, BP neural network has powerful simulation calculation ability, and can approximate any non-linear mapping capabilities[14]. For this reason, it can be used as a suitable tool to generate individual exercise prescription for elderly[13].

## Methods

### Data collection

We collect and summarize previous experimental research which improve the CRF of elderly. PubMed, EBSCO, Web of Science and CNKI were screened for research published in 1989 to 2021(last search in June 2021). A combination of each keyword under the three topics was used as search terms. The topics of this study were older adults, exercise intervention, and cardiorespiratory fitness (Table 1). Synonymous search terms were also added by referring to the retrieved articles during the literature search. Filters were used, including research when written in English or Chinese, designed as randomized controlled trials or self-controlled trials (reviews were excluded) and included elderly aged at least 60 years old were included. Only research with an exercise intervention in older adults, with an outcome focused on VO_2max_ were included; research focusing on the effect of nutritional supplementation was not eligible. Allow research with or without a control group. A total of 63 articles and 2078 subjects were included in result[15–77].

**TABLE 1.**
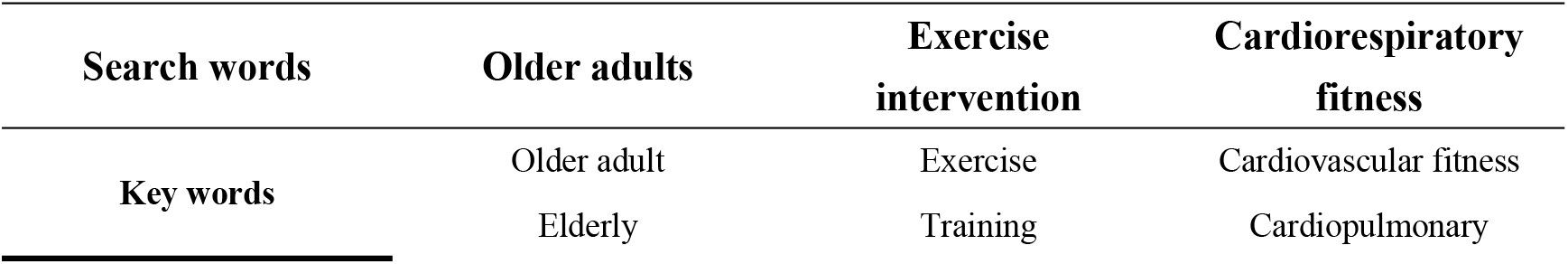
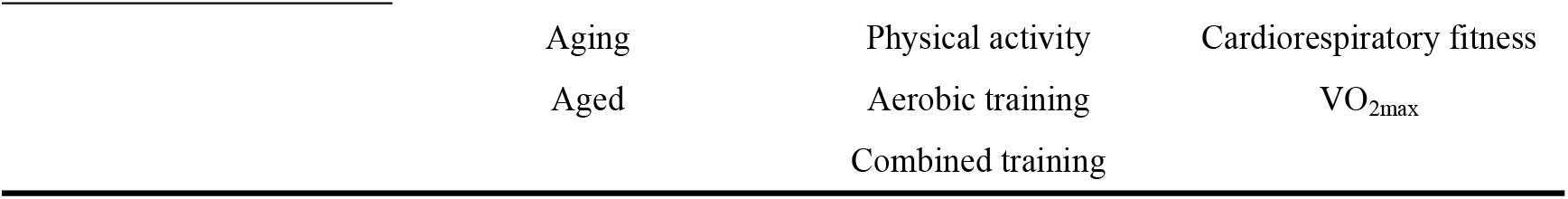
Keywords of dose effect of exercise intervention on improving cardiopulmonary fitness in the elderly.

### Data processing

#### Data encoding process

In each research, we summarize the basic information include age, BMI, VO_2max_ initial value (VO_2max_pre), improvement and sex, and exercise prescription elements of the research’s subjects, include exercise frequency, intensity, time and volume.

To prepare the available data as input and output for training the model, we coded basic information and exercise prescription elements to facilitate loading into the model program. Some information, such as age, BMI and VO_2max_ initial value (VO_2max_pre), are reserve a decimal place. For the setting of VO_2max_ improvement, we use the formula to calculate and reserve two decimal places (VO_2max_ final value = VO_2max_post) and the calculation process is

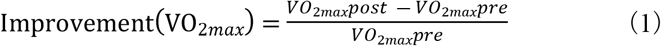

The encoding of the output data set is the same as the input set.

When using the trained BP neural network, not only the input sets should be encoded, but also the output data sets should be encoded. Exercise prescription elements set as our output sets. In the part of exercise intensity set, we extract the average value of the exercise intensity range, convert it to a decimal and subtract 0.5 (we take 50% HRR exercise intensity as zero), such as a study’s exercise intensity is 60%HRR, in our exercise intensity set is 0.1. In the part of exercise time and volume set, we reserve two decimal places.

#### Data expansion process

To expand the sample size reasonably, we expand the collected studies based on the basic information (age, BMI, VO_2max_ initial value) data (mean ± standard deviation) and the number of subjects in each study[78].

The expansion method is implemented on MATLAB 2020a. These generated numbers have passed various statistical tests of randomness and independence, and their calculation can be repeated for testing or diagnostic purposes. The code is as follows:

%% sd1= age’s sd; sd2= BMI’s sd; sd3= VO_2max_ initial value’s sd;

%% mean1= age’s mean; mean2= BMI’s mean; sd3=VO_2max_ initial value’s mean;

a = [sd1 sd2 sd3];

b = [mean1 mean2 mean3];

y = a.*randn(n,1) + b;

Simulate the basic information of each subject in each study, and the information of each subject is generated in the form of a 1*3 matrix. We simulated 2078 groups of sample data in result. By extracting and calculating the above input and output data sets, thus we optimized the input of data in the back propagation neural network.

### BP model

#### Input and output layer settings

The number of neurons in the input unit is directly linked to the prediction outcome. The input data set are the basic information of each sample, and the output date set are the exercise prescription elements of each sample. In order to predict exercise prescription elements at the same time, we construct a multi-input and multi-output model. The input parameters are age, sex, BMI, VO_2max_ initial value and improvement, the output parameters are frequency, intensity, time and volume.

In order to get a precise model, before training the BP neural network, we should set the parameters of the neural network, the selection of neuron transfers functions and neural network training algorithm and errors, etc.[78]. The structure of BPNN in our study is as follows (Fig 1).

**Fig 1.**
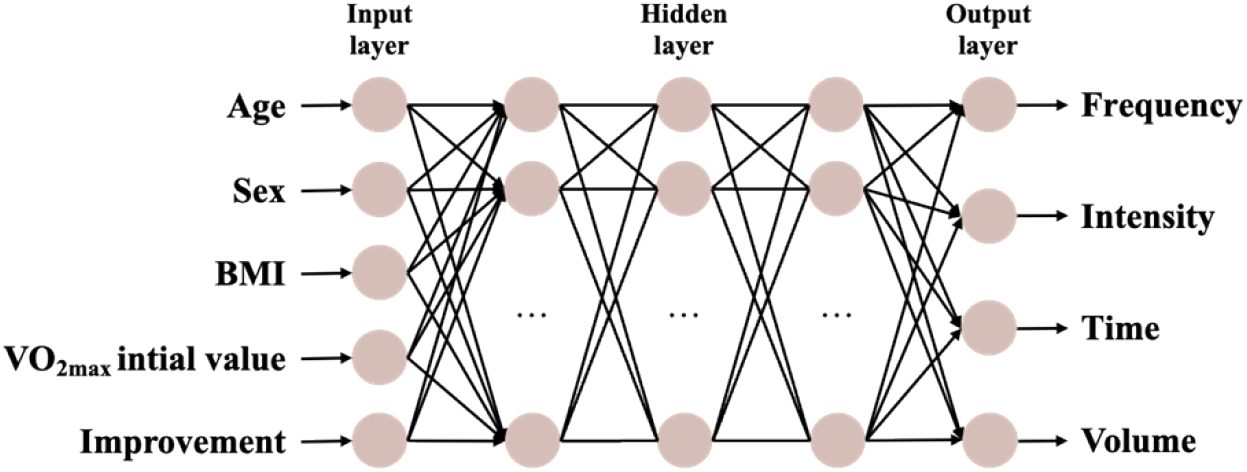
The structure of BPNN.

#### Model algorithm selection

The traditional BPNN algorithm has certain drawbacks, such as slow iterations and a strong tendency to fall into local minima. In our study, the Levenberg-Marquardt (LM) algorithm is used to improve our BPNN algorithm. LM is the fastest algorithm in the MATLAB Deep Learning Toolbox for medium-sized neural networks with fast computational convergence and high accuracy, and is often used to solve fitting and prediction problems, and is highly recommended as the algorithm of choice, although it requires more memory than other algorithms[80].

#### Network layers selection

In BP neural network model, the parameter setup is vital, where the number of nodes in the hidden layer and neurons in the input set unit are the most important variables. It has been proved that if a BP neural network with one hidden layer has enough number of neurons, it can realize any nonlinear mapping. But if the sample number is large, the network with one hidden layer cannot reach an accurate function and the calculation efficiency declines intensively[82]. In order to get an accurate model, we should determine the appropriate number of hidden layers, in which the number of neurons in each hidden layer is determined by (2).

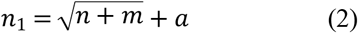

where n_1_ is the hidden neurons, n is the number of input units, m is the number of output units, a is a constant between[83].

According to the empirical formula, the range of the number of neurons is determined as 4-14. In this study, several network architectures with 1, 2, …, 7 hidden layers are tested. We find that, when using our data set, the LM algorithm with three hidden layers and 12/10/8 neurons in layer 1/2/3 performed best in predicting the exercise prescription. The neural network processing is shown in Fig 2.

**Fig 2.**
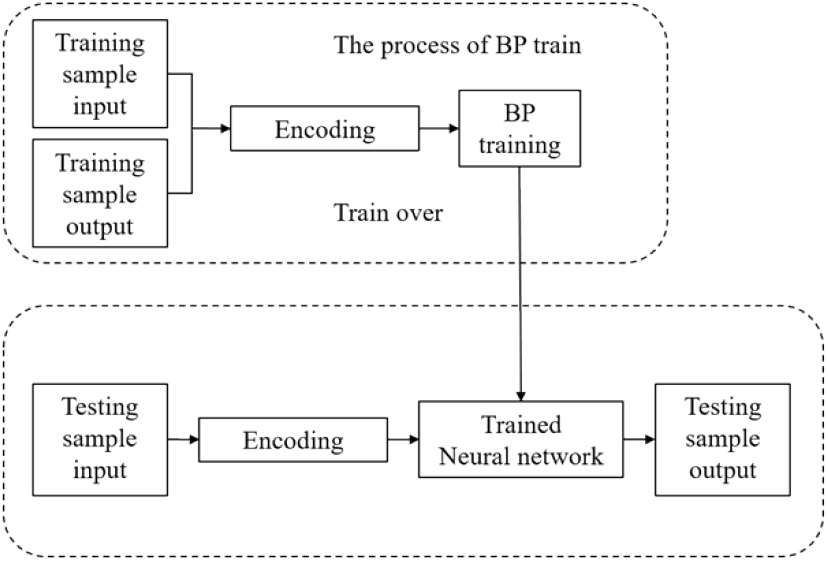
BP neural network training and testing process.

### BP model training and validation

#### BP model training process

The present study needs to determine the appropriate division of the data set to train the model and test the model. Cross-validation is a statistical method that is commonly used in applied machine learning to compare and select a model for a given predictive modeling problem because it is easy to understand, easy to implement, and results in skill estimates that generally have a lower bias than other methods.

The 10-fold cross validation (Fig 3) was done 10 times as an evaluation of the model prediction error by randomly dividing the data set into 10 parts, 9 of which were used as the training set to train the model and the remaining part was used as the test set to test the model error. As a result, 10 test results were obtained, and the error evaluation metrics were averaged to provide an overall measure of the model’s prediction performance.

**Fig 3.**
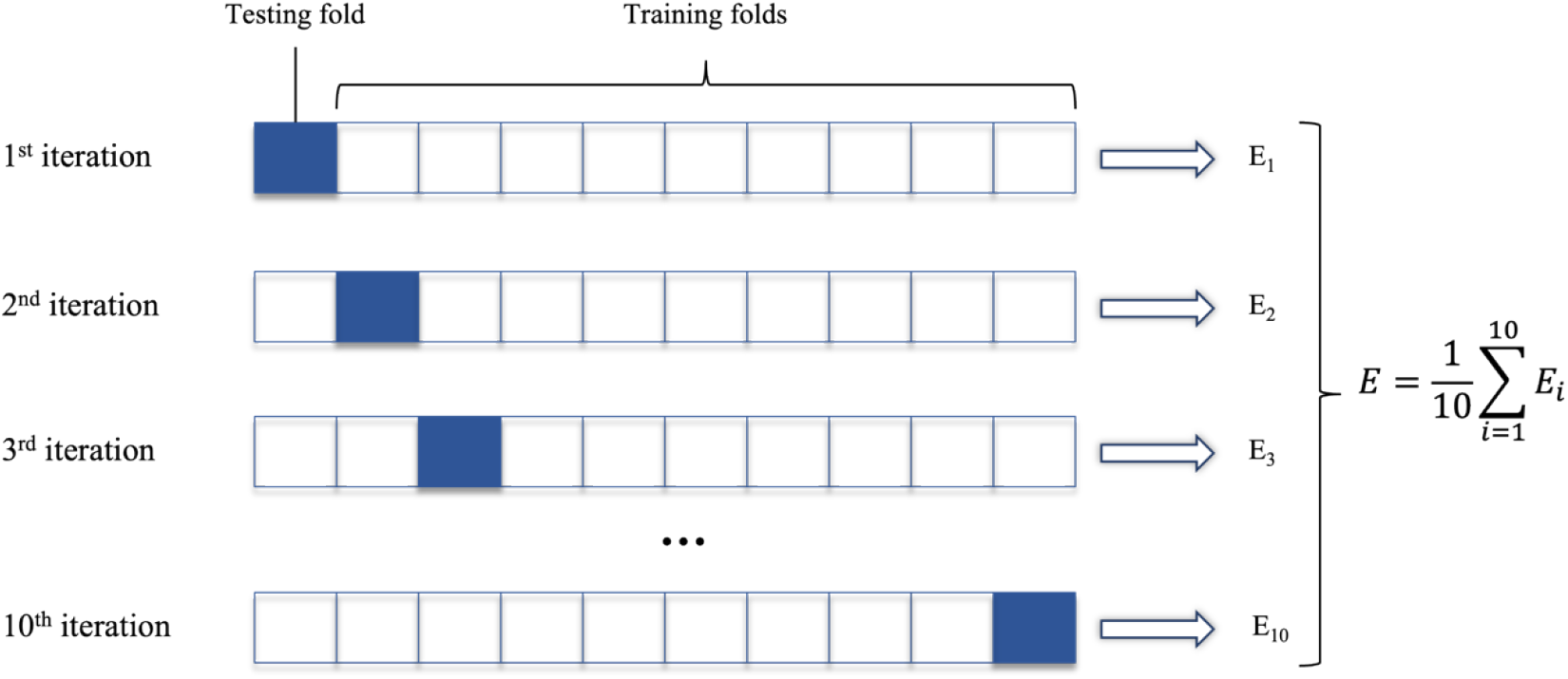
10-fold Cross-Validation.

#### BP model validation

##### (1) Evaluation Indicators

Three evaluation indicators, Mean Absolute Error (MAE), Root Mean Squared Error (RMSE) and R-squared (R^2^) are selected to evaluate the accuracy of the exercise prescription prediction model. Their expressions are as follows:

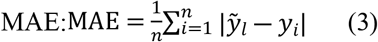

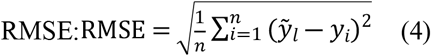

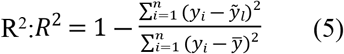

where 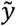 represents the predicted value and y represents the actual value.

##### (2) Instance Verification

Twenty-six elderly female subjects aged 60-79 are recruited to test the practical application effect of the model. The inclusion criteria of the subjects were in accordance with the inclusion criteria of the data set literature. The basic characteristics of the subjects are shown in Table 3. The subjects’ VO_2max_ was estimated from the 6-minute walk performance (r=0.81, SEE=2.68)[84]. The expected improvement was set at 10%. Based on the basic information of these subjects, we get personalized exercise prescription (frequency, intensity, time and volume) of each subject from the model. And they follow the prescription to do aerobic exercise. Heart rate monitors (Polar Electro, H10) can record a person’s heart rate during exercise, allowing them to determine and adjust the intensity of their workout.

**Table 3.** The basic characteristics of the subjects.

| Basic characteristic |  |
| --- | --- |
| Age (years) | 67.1±4.8 |
| Sex | Female |
| BMI (kg/m<sup>2</sup>) | 24.5±3.7 |
| Pre $\text{VO}_{2\max}$ (mL kg<sup>-1</sup>min<sup>-1</sup>) | 12.4±2.2 |
| Improvement (%) | 10 |

## Result

### Neural network design

We selected 63 articles and 2078 datasets as the dataset. After dividing the dataset into 10 randomly, 9 of them (N=1870) are selected as the training set for building the model, and the remaining one (N=208) is used as the testing set for the model. The framework structure of the model is 5 dimensions for the input layer, 12-10-8 dimensions for the 3 hidden layers, and 4 dimensions for the output layer. The results of each round of cross-validation are shown in Table 4.

**Table 4.** Comparison of error indicators of each round of cross-validation.

| Round | RMSE | MAE | $R^2$ |
| --- | --- | --- | --- |
| 1 | 1.4190 | 1.6433 | 0.9946 |
| 2 | 1.3964 | 1.4312 | 0.9947 |
| 3 | 1.4777 | 1.6964 | 0.9941 |
| 4 | 1.6234 | 1.3363 | 0.9930 |
| 5 | 1.5250 | 1.4587 | 0.9938 |
| 6 | 1.3549 | 1.4226 | 0.9951 |
| 7 | 1.2620 | 1.2711 | 0.9957 |
| 8 | 1.5960 | 1.8182 | 0.9932 |
| 9 | 1.2366 | 1.5394 | 0.9959 |
| 10 | 1.4918 | 1.5887 | 0.9940 |
| AVG | 1.4383 | 1.5206 | 0.9944 |
| Best | 1.2366 | 1.5394 | 0.9959 |

From the error indicators in Table 4, the average and best of RMSE of this BP neural network are 1.4383 and 1.2366. Similarly, the average and best of MAE of this network are 1.5394 and 1.5206. According to previous study, in their intelligent physical health evaluation model, the MAE of the BPNN is about 2.649 and the RMSE of the BPNN is about 3.032 [98]. Likewise, in another study, in their blood pressure prediction with BPNN, their MAE of diastolic blood pressure (DBP) is about 2.43, and systolic blood pressure (SBP) is about 3.97. And their RMSE of DBP and SBP are about 4.18 and 8.9[85]. Besides, in our study, average R^2^ are all above 0.99, which prove that our BP neural network have good results as elderly exercise prescription elements prediction model. Fig 4(a) shows the training error diagram of the LM-BPNN. The horizontal axis shows the training epoch, and the vertical axis shows the mean squared error (MSE) on the data set. The best training performance of the model is 4.5416 at epoch 47. Fig 4(b) shows the regression fitting values R for the training set, validation set, testing set and the overall are 0.99659, 0.99408, 0.99326 and 0.99572 respectively. The R values measure the correlation between outputs and targets. An R value of 1 means a close relationship, 0 a random relationship.

**Fig 4.**
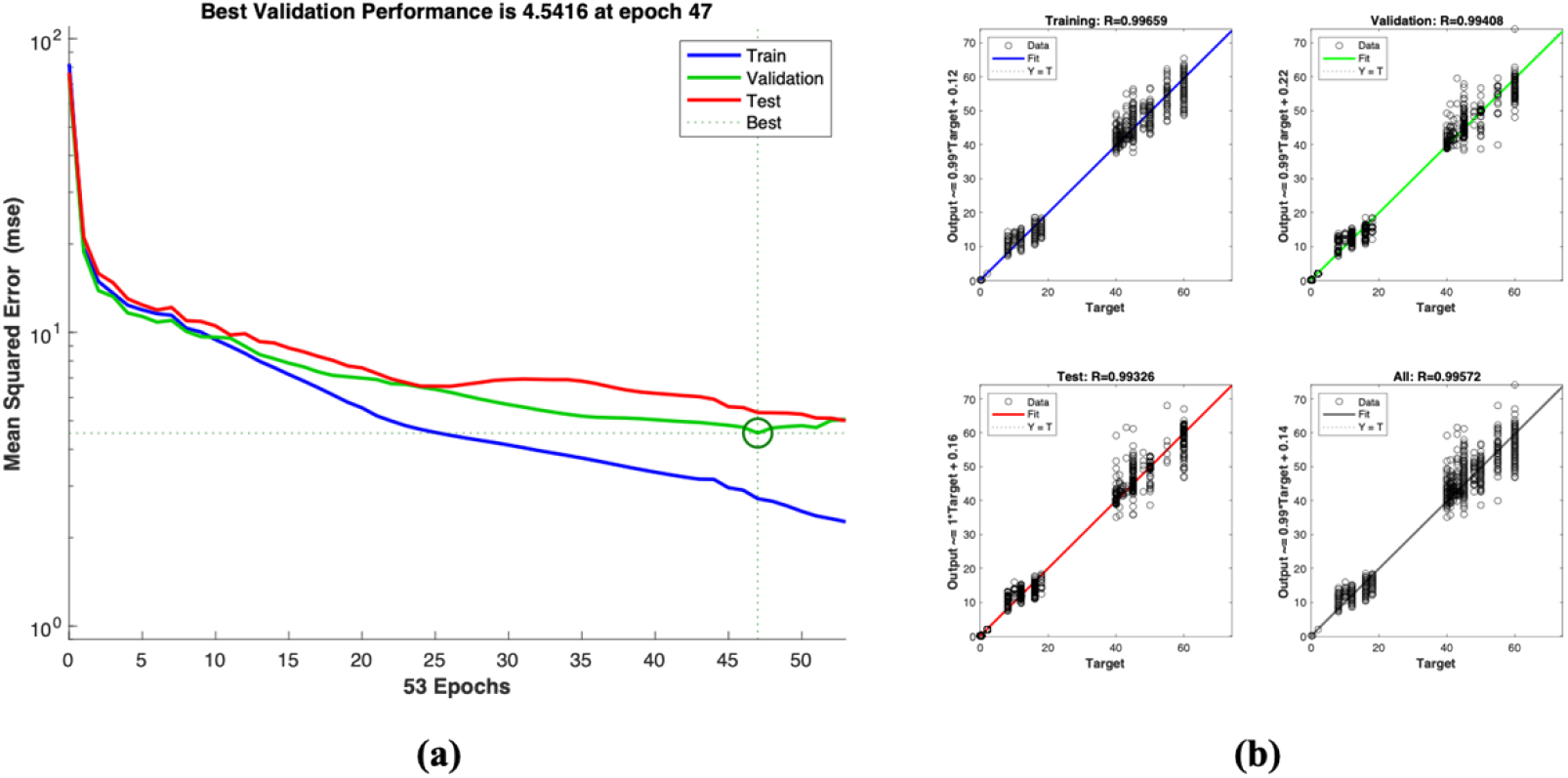
Model training error situation. (a) Fitting regression diagram of LM-BPNN model; (b) Training error diagram of LM-BPNN

Figs 5-7 show the predicted result of the BP neural network on best test round. These figures show the predicted results between the true value and prediction value, we can see that a small difference between the prediction value and true value and demonstrates a better prediction trend and accuracy in exercise time and period. For the prediction of exercise intensity, our model can also make a basically accurate prediction.

**Fig 5.**
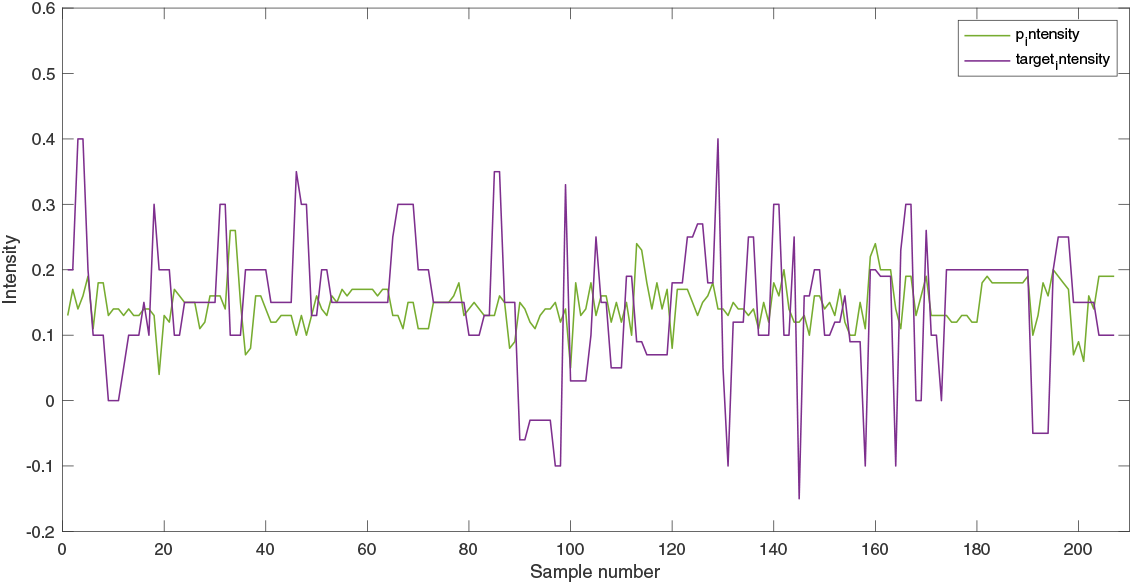
Comparison between the true results and BPNN prediction results at Intensity.

**Figure 6.**
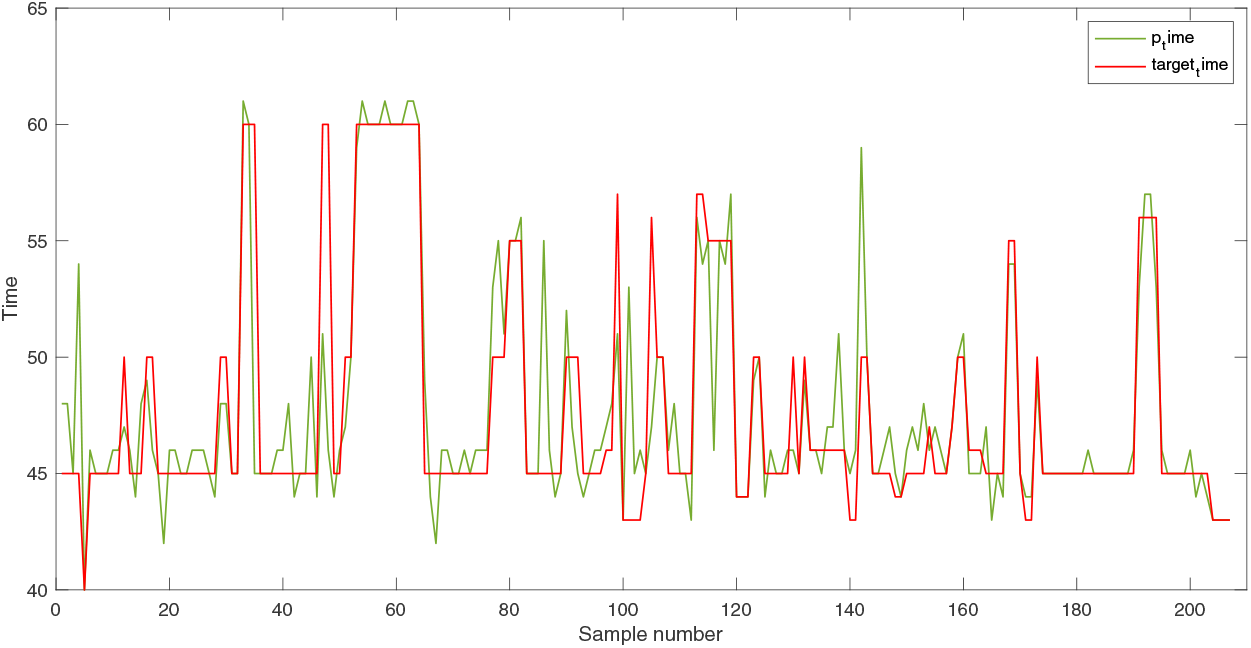
Comparison between the true results and BPNN prediction results at Time.

**Figure 7.**
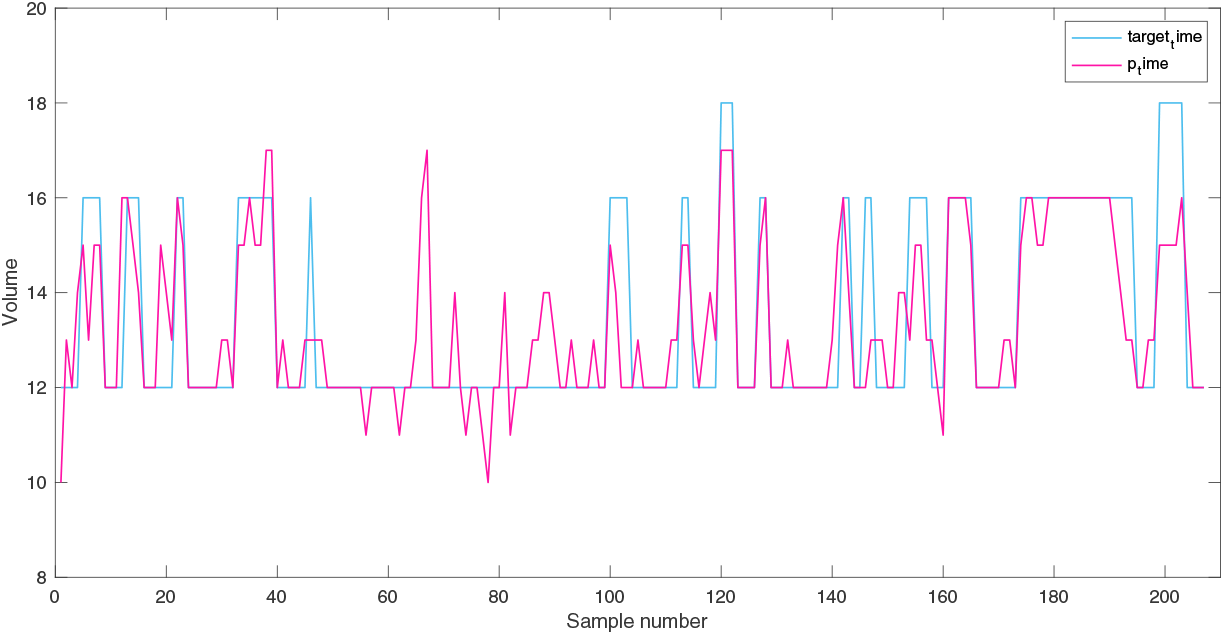
Comparison between the true results and BPNN prediction results at Volume.

### Instance Verification results

The exercise prescriptions generated from the model are shown in Table 5. It can be seen that the frequency of exercise is 2 times per week, the exercise intensity is mostly concentrated at 64% HRR, the average exercise time is about 48 minutes (range from 45 minutes to 55 minutes), and the volume is around 12 weeks.

**Table 5.** Model-generated exercise prescriptions for subjects.

|  | Frequency (d/w) | Intensity (%HRR) | Time (min) | Volume (w) |
| --- | --- | --- | --- | --- |
| **M±s** | 2.0±0 | 64.2±2.7 | 48±3 | 11.9±0.6 |
| **Range** | 2 | 59.9-67.4 | 45-55 | 10.9-12.8 |

After finish their exercise intervention,the 6MWT is measured for estimating VO_2max_ again. The differences in pre VO_2max_, post VO_2max_ and the expected improvement in VO_2max_ are shown in Fig 8. It can be seen that the post VO_2max_ was significantly different from the pre VO_2max_ and improved by 10.1%. There was no difference between the post VO_2max_ and the expected improved VO_2max_, indicating that the actual improvement in the model was largely in line with the expected improvement.

**Fig 8.**
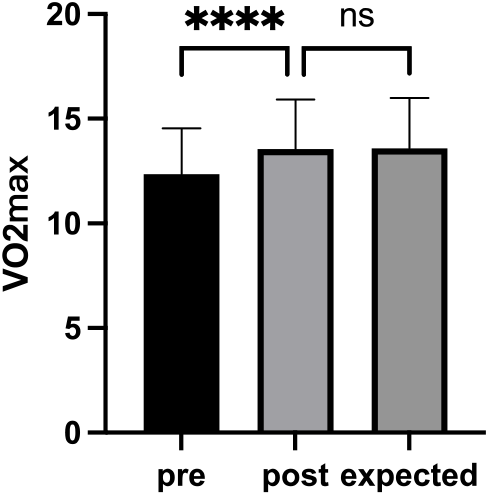
Difference between pre VO_2max_, post VO_2max_ and expected improvement in VO_2max_. Note: ****, p<0.0001; ns: no significant.

Table 6 shows the “hit rate” (also called predictive accuracy) of CRF improvement for 26 subjects over a range of one standard deviation and a range of 1.96 times standard deviations[86,87].A total of 20 subjects improved within one standard deviation and 25 subjects improved within two standard deviations, indicating that each subject had good CRF improvement.

**Table 6.** “Hit rate” of improvement at one standard deviation and 1.96 times standard deviation range.

|  | Range | Hit rate |
| --- | --- | --- |
| **M±sd** | 1.9%-18.2% | 76.9% |
| **M±1.96sd** | -6.2%-26.3% | 96.2% |

### Limitations and further work

Compared to previous studies, we innovatively use the BP neural network model to recommend exercise prescriptions for elderly. Although the model can achieve good results, it still has limitations. In our study, the reason why exercise intensity is not well predicted is mainly due to the variety of exercise intensity, such as percentage of heart rate reserve, percentage of VO_2max_, and percentage of HR_max_, they designed for elderly in each research we collected, so when we process each exercise intensity from each study, some random errors will generate. Therefore, the exercise intensity prediction of this model cannot achieve the general accuracy of predicting exercise time and exercise volume.

However, in the reality exercise intervention for elderly, they do not exercise at a fixed exercise intensity, but exercise within a range. Likewise, when we encode the exercise intensity set, we use the middle value of the exercise intensity range in each study, so the predicted exercise intensity result can also be used as a reference median value. We can use an increase and decrease of 5% to 10% in the median as a range of predicted exercise intensity.

Although our model can achieve a good degree of simulation, it is currently not possible to give different prescriptions for each independent individual. For example, if two elderly people have the same parameters, the prescriptions generated from the model will be the same. So the actual improvement effect still needs to be verified in practice.

In the following, we can consider expanding the sample size and research parameters of the model, adding a new algorithm to pre-process the exercise intensity set and then inputting it into the BP neural network for prediction, and use this model as the core to establish an application that can be used for elderly.

## Conclusion

In this study, we propose to use a model based on BP neural network to create individualized CRF exercise prescriptions for elderly. The model consists of 5 layers BP neural network and the structure is 5-12-10-8-4. By inputting the basic information of elderly after exercise evaluation and the expected improvement degree of CRF, this model can generate a suitable exercise prescription. After expanding the collected data, an evaluation system for evaluating the sample cardiopulmonary health (VO_2max_) is obtained through machine learning and modeling, which provides a convenient solution for future elderly health exercise. The experimental results show that the exercise prescriptions predicted by our proposed model are closely related to the actual exercise prescriptions. To examine the actual improvement of the model, we verified the improvement in CRF by recruiting subjects to perform the model-generated exercise prescriptions for the intervention. At the end of the intervention, the post-intervention VO_2max_ was close to the expected improved VO_2max_, which can indicate the high validity and reliability of the exercise prescription model based on BP neural network for the elderly.

This work demonstrates the potential of the BP neural network model in predicting individualized sports for elderly. This article introduces our method from several parts, including data collection, data expansion and the model establishment based on BP neural network. The advantages of the BPNN model come from the capacity to learn input data at multiple spatial and temporal levels of abstraction. In summary, our study proposed in this paper provides an innovative and effective solution for the health exercise of elderly.

## Data Availability

All elderly dataset and model files are available from the https://github.com/reallycolin/BPNN-for-elderly-exercise-prescription

## Acknowledgments

Funding was provided by the National Key R&D Program of China (2018YFC2000604).

## Institutional Review Board Statement

The research proposal was approved by the Institutional Review Board of Beijing Sport University (BSU IRB) and all participants gave written informed consent prior to study participation (No. 2018018H).

## Author contributions

**Conceptualization:** Chunyan Xu, Yiran Xiao, Lantian Zhang, Xiaozhen Ding.

**Data curation:** Yiran Xiao, Lantian Zhang, Xiaozhen Ding.

**Formal analysis:** Yiran Xiao, Xiaozhen Ding.

**Funding acquisition:** Chunyan Xu.

**Investigation:** Chunyan Xu, Yiran Xiao, Lantian Zhang, Xiaozhen Ding.

**Methodology:** Chunyan Xu, Yiran Xiao, Lantian Zhang, Xiaozhen Ding.

**Project administration:** Chunyan Xu.

**Resources:** Chunyan Xu, Yiran Xiao, Lantian Zhang, Xiaozhen Ding.

**Software:** Chunyan Xu, Yiran Xiao, Lantian Zhang.

**Supervision:** Chunyan Xu.

**Validation:** Chunyan Xu, Yiran Xiao, Lantian Zhang, Xiaozhen Ding.

**Visualization:** Yiran Xiao, Lantian Zhang, Xiaozhen Ding.

**Writing – original draft:** Yiran Xiao, Lantian Zhang, Xiaozhen Ding.

**Writing – review & editing:** Chunyan Xu, Yiran Xiao, Lantian Zhang, Xiaozhen Ding.

## Supporting information

